# Prevalence of multimorbidity in migrants, refugees, asylum seekers and displaced persons and association with mortality: A systematic review protocol

**DOI:** 10.1101/2022.09.02.22279507

**Authors:** A Abrahamsson, A Rozalen Gonzalez, BI Nicholl, CA O’Donnell, FS Mair

## Abstract

**Background:** Multimorbidity is the presence of two or more physical and/or mental long-term health conditions and is associated with aging, but also with socio-economic deprivation. Immigration status contributes to socio-economic deprivation, potentially increasing prevalence of multimorbidity. Prevalence of multimorbidity in different migrant populations, such as asylum seekers, refugees and displaced persons, and its effect on mortality is poorly understood and no systematic review on the topic exists to date.

**Objective:** To assess what is known about the prevalence of multimorbidity in refugees, asylum seekers, migrants and displaced persons and about the association, if any, between multimorbidity and mortality in these populations.

**Design:** Systematic review of the literature. The following electronic medical databases will be searched: MEDLINE, Embase, Scopus, CINAHL and The Cochrane Library. A narrative synthesis of findings will be undertaken, and meta-analysis considered if appropriate. Title, abstract and full paper screening will be undertaken independently by two reviewers. Studies will be limited to English and Spanish language and publication date from 2000 onward. The Newcastle-Ottawa Scale for Cohort Studies will be used as a quality assessment tool. This protocol adheres to the Preferred Reporting Items for Systematic Reviews and Meta-Analyses Protocols (PRISMA) 2020 guidelines.

**Conclusion:** Understanding the prevalence of multimorbidity amongst refugees, migrants, asylum seekers and displaced persons, and its association with mortality will provide valuable insights to inform practice and policy responses to increasing global migration.

## Introduction

### Rationale

According to the World Health Organisation, chronic diseases are responsible for 71% of deaths globally.^1^ The burden of chronic diseases, also referred to as non-communicable diseases, is a concern from a global public health perspective partly because of its high prevalence and partly because it is often complex, with multimorbidity (presence of two or more long-term health conditions^2^) being increasingly common. Subsequently, a holistic approach needs to be taken when dealing with multimorbidity, to account for and promote the understanding of the interactions between physical, mental and social conditions.^3^ Although the prevalence of multimorbidity tends to be associated with increasing age, there is evidence that social deprivation is also strongly correlated with multimorbidity.^4-6^ Migrants, refugees, asylum seekers and other displaced populations are acknowledged to be at higher risk for chronic illness because of high levels of poverty and homelessness secondary to social, political or economic exclusion^7^ and therefore may be at higher risk of multimorbidity and adverse health outcomes.

Migration can be understood as a population’s movement from one geographical area to another for an indefinite period for diverse circumstances.^8^ This wide definition includes voluntary types of migration for example work employment or family reunification. Forced migration refers to “a migratory movement which, although drivers can be diverse, involves force, compulsion or coercion”.^9^ Types of forced migration include refugees, asylum seekers and displaced persons. According to the United Nations High Commissioner for Refugees (UNHCR), a refugee is someone who “is unable or is unwilling to return to their country of origin owing to a well-founded fear of being persecuted for reasons of race, religion, nationality, membership of a particular social group, or political opinion”.^10^ Asylum seeker is a general term for “any person who is seeking international protection. In some countries, it is used as a legal term referring to a person who has applied for refugee status or a complementary international protection status and has not yet received a final decision on their claim. It can also refer to a person who has not yet submitted an application but may intend to do so, or may be in need of international protection”.^11^ Displaced person is defined as “Persons or groups of persons who have been forced or obliged to flee or to leave their homes or places of habitual residence, either across an international border or within a State, in particular as a result of or in order to avoid the effects of armed conflict, situations of generalized violence, violations of human rights or natural or human-made disasters.”^9^ Undocumented migrant, or migrant in an irregular situation is a term for “a migrant who, owing to unauthorized entry, breach of a condition of entry, expiry of a visa or stay permit, or failure to comply with an expulsion order, has no legal permission to stay in a host country.^11^

Particular challenges in this group of people may be poor health literacy and negative experience with accessing healthcare, among others.^12,13^ The process and experience of forced migration also places individuals in circumstances that can affect their physical and mental wellbeing, making them vulnerable to illness and deterioration of existing long-term conditions.^14,15^ Migration is a growing issue internationally with numbers of global migrants growing faster than the global population, rising from 2.8% in 2000 to 3.6% in 2020.^16^

For host countries to be better equipped to respond to the needs of migrant populations, especially those with existing long-term conditions and multimorbidity, a better understanding of the prevalence of multimorbidity and impacts in migrant populations is needed. However, there has as yet been no systematic literature review exploring the prevalence of multimorbidity in migrant groups, or the relationship between multimorbidity and mortality in these population groups. Examining such relationships will give health professionals a better understanding of the health challenges faced by migrant populations and help inform future health service delivery for these groups.

### Objectives

This systematic review will assess what is known about the prevalence of multimorbidity in refugees, asylum seekers, migrants and displaced persons and about the relationship, if any, between multimorbidity and mortality in these populations

## Methods

The Preferred Reporting Items for Systematic Review and Meta-Analysis Protocols (PRISMA-P) 2020 reporting guidelines were used to prepare this protocol.^17^

### Eligibility criteria

Inclusion and exclusion criteria are summarised in Table 1.

**Table 1.** Inclusion and exclusion criteria.

| Inclusion Criteria |  |
| --- | --- |
| Population | Migrants (i.e., a person who has resided in a foreign country for any length of time irrespective of the reasons for the move)<sup>8</sup>, refugees (i.e., people who had to flee their country of origin for various |
|  | reasons beyond their control and seek protection in another country)<sup>10</sup>, asylum seekers (i.e., Asylum seeker is a person who applies for recognition of refugee status and whose application is in the process of being assessed)<sup>11</sup>, displaced persons (i.e., a person who has been forced to leave his or her home or habitual residence, either across an international border or within a State)<sup>9</sup>, and undocumented migrants (migrant who owing to e.g. expiry of visa or entry permit have no legal permission to stay in the country)<sup>11</sup> with multimorbidity.<br>Adults (individuals aged $\geq 18$ years); however, if the study included <18-year-olds, it may still be included if multimorbidity prevalence is reported separately for adults and children. |
| **Exposure** | Multimorbidity ($\geq 2$ physical and/or mental health conditions) which may include:<br>○ a list of chronic diseases from different sources (e.g., health records, census, administrative data, or prescription records)<br>○ self-reported (e.g., interview or survey) |
| **Measurement of exposure** | Multimorbidity condition count including simple chronic disease count or the use of specific measures (e.g., Charlson Comorbidity Index). |
| **Comparator** | Migrants, refugees, asylum seekers, and displaced persons without multimorbidity.<br>Individuals in the native host country population with or without multimorbidity.<br>Individuals in the country of origin with or without multimorbidity.<br>Or no comparator |
| **Outcome** | Prevalence of multimorbidity and, if mentioned, mortality rates related to multimorbidity. |
| **Settings** | Any (e.g., primary, or secondary care, refugee camps, community settings) |
|  | Publications dated between 2000 and 2022.<br>Studies from any location worldwide.<br>English and Spanish language.<br>Peer-reviewed journals with full-text available.<br>Quantitative longitudinal (retrospective and prospective), cross-sectional or observational studies. |
| **Exclusion Criteria** |  |
| **Population** | Studies focusing solely on paediatric (<18 years) or animal populations<br>Non-migrant population, non-refugee, non-displaced persons, non-asylum seekers populations |
| **Exposure** | Studies focusing only on single or two conditions/ comorbidity |
| **Outcome** | Non-chronic short-term diseases or acute diseases |
| **Types of studies** | Qualitative studies<br>Non-English or Spanish language<br>Grey literature, non-peer reviewed journals, dissertations, academic theses, case reports, conference abstracts or books<br>Duplicated studies |

#### Study design

Empirical studies using quantitative methods will be eligible for this review. We will accept longitudinal (prospective or retrospective), cross-sectional and observational studies conducted anywhere in the world. We will accept studies conducted in any form of setting, e.g. primary or secondary care, refugee camps or community settings. Qualitative studies will be excluded, as will be grey literature, non-peer reviewed articles, dissertations, academic papers, case reports/series, conference abstracts, systematic reviews and books. Publication date restrictions will be from 2000 onwards. These time restrictions have been chosen as the term multimorbidity was only introduced as a Medical Subject Heading (MESH) in 2018^18^ and the literature on multimorbidity only began to emerge after 2000 with even comorbidity only becoming a MESH term in 1990.^19^ Also, migration has only been noted by international agencies to become a particularly important issue since the start of the 21^st^ century.^20,21^ The timeframe therefore intends to avoid collecting too many irrelevant studies.

#### Population

Participants of both sexes and with any migrant status (e.g. migrants, refugees, asylum seekers and displaced persons) of any ethnicity and from any origin country in the world with multimorbidity (≥ 2 long-term non-communicable health conditions) will be included. The study must focus on adults (≥ 18 years); but if younger age groups are processed in the same study they will be included for this review if data on the population can be separated or distinguished separately

#### Exposure

The exposure of interest is the presence of two or more long-term conditions in adult refugees, migrants, asylum seekers, immigrants or displaced persons. Multimorbidity assessed by any numerical count or validated measure (e.g. Charlson Comorbidity Index^22^) will be accepted, including lists of chronic diseases obtained from sources such as health records, national administrative data, census or prescription records, as well as self-reported data from interviews or surveys. Studies that focused only on a single or two conditions will be excluded. Also excluded will be studies that focused solely on acute infections or short-term diseases.

The other outcome of interest will be mortality associated with multimorbidity among migrants, refugees, asylum seekers and displaced persons.

#### Comparators

Studies where the comparator was either migrants, refugees, asylum seekers, or displaced persons not having multimorbidity (≥2 long term conditions), individuals in the country of origin with or without multimorbidity, or the local native population in the new host country with or without multimorbidity will be included. However, studies that did not include a comparator/control group will not be excluded.

#### Outcomes

The outcomes of interest will be the prevalence of multimorbidity, and mortality associated with multimorbidity. Studies which provide data on either, or both, of these outcomes will be included.

#### Publication type

Studies must be full-text, fully accessible, published peer-reviewed articles. Conference abstracts, case reports/series, dissertations/theses, books, systematic reviews and editorials/commentaries/letters will be excluded.

#### Language

Studies must be in the English or Spanish language.

### Information sources

The following electronic databases will be searched: MEDLINE (Ovid), Embase (Ovid), CINAHL (EBSCOhost), The Cochrane Library (Wiley) and Scopus (Elsevier). The International Prospective Register of Systematic Reviews (PROSPERO) will be monitored for other ongoing and completed systematic reviews on multimorbidity prevalence in migrants, refugees, asylum seekers and displaced persons.

### Search strategy

This review will follow the Population, Exposure, Control/Comparator, Outcomes, Settings and Study design (PECOS) framework to design the search strategy.^23^ The search strategy will combine the following key concepts: (1) Migrant/Refugees/Asylum seekers/Displaced persons, (2) multimorbidity and (3) prevalence/mortality, and will use a varied set of search index terms and keywords based on the elements of the PECOS framework. The full search strategy can be found in the supplementary material. Each database will be searched individually with the search strategy adapted to reflect the differing subject index terms used by each database. Boolean operators will be combined with advanced search features such as multi-field search, operators, truncation/wildcards and limits, to assure appropriate width of the search.

### Study records

#### Data management

The results of the literature search will be downloaded to a reference management software for systematic reviews and duplicates removed. Thereafter double screening will be undertaken.

#### Selection process

The process of selecting eligible studies will be conducted in two stages, title/abstract screening, and full-text screening. Title and abstracts of studies identified will be independently screened by ARG and AA, against predefined eligibility criteria. Studies that meet the predefined eligibility criteria will undergo full-text screening. Where there are conflicting views on titles or abstracts, they will undergo full-text screening. Full-text articles will be obtained and independently reviewed by ARG and AA, again against the predefined eligibility criteria. Disagreements at this stage will be resolved by discussion and/or consultation with a third reviewer (FSM or BN). A reference search and citation search will also be performed on included full-text articles to identify any additional relevant studies.

#### Data collection process

Data from studies included after full-text screening will be extracted and recorded in a predefined data extraction table designed by ARG. The data extraction will be guided by similar systematic reviews on the theme of multimorbidity.^24-26^ The extraction items included will be appropriately adapted to be relevant to the aims and objectives of this systematic review. The data extraction table will be completed by two reviewers (ARG, AA) and discrepancies will be resolved by discussion and/or consultation with a third reviewer (FSM or BN).

### Data items

#### Study characteristics

Details relating to study design, country of study and data collection period will be extracted. In addition first author name and year of publication will be extracted.

#### Population

Study population characteristics, such as age, sex and ethnicity, type of migrant, as well as sample size and response rate will be extracted. Any available information regarding lifestyle, such as rates of smoking, physical activity and alcohol consumption will be collected.

#### Exposure

The way multimorbidity is defined and measured will be recorded (e.g. Charlson Comorbidity Index^22^ or other score or chronic disease(s) count) and if a multimorbidity count was provided, what conditions it included and how many of them will be recorded. Details on prevalence and patterns of multimorbidity will also be extracted. Any available details on physiological measurements related to chronic conditions, such as BMI, blood pressure, glucose/HbA1c etc, will also be extracted.

#### Comparator

Comparators, if any, described and reported for each study will be recorded.

#### Outcome

The results of each included study will be recorded. Details about the prevalence of multimorbidity and mortality, including information on the extent of multimorbidity (i.e. chronic disease(s) count) will be extracted. For studies including information on associated mortality, mortality hazard ratios will also be extracted.

### Outcome and prioritisation

The primary outcome of interest is prevalence of multimorbidity in migrants, refugees, asylum seekers and displaced persons. Any measurement of multimorbidity will be accepted, from simple chronic disease count to specific tools, such as the Charlson Comorbidity Index.^22^ Of equal priority and interest is the relationship between multimorbidity and mortality in these groups.

### Risk of bias in individual studies

The Newcastle-Ottawa Scale (NOS) for Cohort Studies will be used as the quality assessment tool of choice.^29^ The NOS scale will be used independently to assess risk of bias in included studies in this systematic review. This scale uses a star-rating system to appraise three main areas of the cohort studies: (1*) the selection process*, (2) *comparability of the cohorts on the basis of the design or analysis* and (3) *the outcome*. A maximum of nine stars can be awarded.^27^ The NOS scale can also be adapted and may be for this review, following the example of other systematic reviews such as Chiang et. al.^26^ Disagreements between reviewers at this stage will be discussed and resolved by a third reviewer (FSM or BN), if required.

### Data synthesis

If applicable, meta-analysis will be undertaken. However, as we expect heterogeneity in the literature in terms of how multimorbidity is defined, and in how outcomes are reported, a narrative synthesis is more likely to take place. A narrative synthesis of findings will describe the following from each study: study characteristics, population, exposure, comparator, outcomes, risk of bias (quality) assessment and study inconsistencies, if any. If subgroup analysis by migrant group is possible, this will be undertaken.

### Ethics approval and dissemination

As this systematic review will not contain individual patient data and is using existing published data, ethics approval is not required. The results of this review will be disseminated via relevant scientific conferences, peer-reviewed journals and social media.

## Discussion

This systematic review will synthesise the current existing literature to determine what is known about the prevalence of multimorbidity in migrant, refugee, asylum seeker and displaced persons. It will also synthesise what is known about multimorbidity and mortality in these groups. The prevalence of multimorbidity may be higher in the various migrant populations, due to the various additional challenges faced by such individuals,^12,13^ or prevalence may be lower as migrants tend to be young and healthier than the native population in their new host country, sometimes referred to as the “healthy immigrant effect”.^28,29^ To our knowledge, this systematic review will be the first to examine the current information available about the prevalence of multimorbidity in various migrant groups and will therefore identify gaps in knowledge about migrant health and contribute to overall understanding in this area.

Strengths of this systematic review will be the adherence to the PRISMA-P 2020 guidelines to ensure clarity and transparency in reporting.^17^ Furthermore, all screening and quality appraisal will be performed by two reviewers independently with a third reviewer adjudicating disagreements. A limitation of this study is that the definition of multimorbidity and reporting of outcome in the literature are expected to vary considerably. As is the terminology used when describing different migrant groups, and the intended understanding of the terms used in relation to migrants. Due to this complexity and heterogeneity, meta-analysis may not be possible which may be a limitation. The search strategy will also be restricted to English and Spanish language studies, and the period 2000 to 2022.

We expect the review to provide new insights on this topic that will highlight areas requiring further research as well as providing evidence to inform future clinical practice and policy.

## Data Availability

Data produced in the present study are available upon reasonable request to the authors after publication of the full paper on this systematic review

## Acknowledgement

We want to acknowledge Sonny Maley, librarian at the University of Glasgow’s Medical Veterinary and Life Science (MVLS) College, for his assistance in developing the search strategy for this review.

## Author contributions

ARG and FSM conceived the study and developed the search strategy, inclusion/exclusion criteria and data extraction table. The protocol was drafted by AA and ARG with input from FSM, BN and COD. FSM and BN will act as third reviewers when needed. All authors have approved the final manuscript for submission.

## Supplementary Data

### Search strategy

#### Search strategy CINAHL (EBSCOhost)

| # | Searches (All text without suggested subject terms) |
| --- | --- |
| 1 | (MH "Comorbidity") |
| 2 | (MH "Chronic Disease+") |
| 3 | (MH "Prevalence") |
| 4 | (MH "Death+") |
| 5 | (MH "Mortality+") |
| 6 | (MH "Refugees+") OR (MH "Immigrants+") OR (MH "Emigration and Immigration") OR (MH "Undocumented Immigrants") OR (MH "Transients and Migrants") OR (MH "Refugee Camps") |
| 7 | S1 OR S2 |
| 8 | S3 OR S4 OR S5 |
| 9 | S6 AND S7 AND S8 |

#### Search strategy Cochrane library

| # | Searches |
| --- | --- |
| 1 | MeSH descriptor: [Multimorbidity] explode all trees |
| 2 | MeSH descriptor: [Comorbidity] explode all trees |
| 3 | MeSH descriptor: [Prevalence] explode all trees |
| 4 | MeSH descriptor: [Refugees] explode all trees |
| 5 | MeSH descriptor: [Emigrants and Immigrant] explode all trees |
| 6 | MeSH descriptor: [Transients and migrants] explode all trees |
| 7 | #1 OR #2 |
| 8 | #4 OR #5 OR #6 |
| 9 | #7 AND #3 AND #8 |

#### Search strategy EMBASE

| # | Searches |
| --- | --- |
| 1 | exp multiple chronic diseases/ |
| 2 | exp comorbidity/ |
| 3 | (multimorbid* or "multi morbid*" or comorbid* or "co morbid*").ti,ab. |
| 4 | (condition* adj3 count*).ti,ab. |
| 5 | ("multi condition*" or "multi disease*" or "multi disorder*" or multicondition* or multidisorder* or multidisease*).ti,ab. |
| 6 | 1 or 2 or 3 or 4 or 5 |
| 7 | exp migrant/ |
| 8 | exp refugee camp/ or exp refugee/ |
| 9 | (migrant* or refugee* or immigrant* or emigrant*).ti,ab. |
| 10 | (displace* adj3 person*).ti,ab. |
| 11 | (asylum* adj3 seeker*).ti,ab. |
| 12 | 7 or 8 or 9 or 10 or 11 |
| 13 | exp Mortality/ |
| **14** | exp Death/ |
| **15** | exp prevalence/ |
| **16** | ((death* or fatalit* or mortalit*) adj3 (rate* or declin* or determinant* or "differential exces*")).ti,ab. |
| **17** | prevalence*.ti,ab. |
| **18** | 13 or 14 or 15 or 16 or 17 |
| **19** | 6 and 12 and 18 |
| **20** | limit 19 to english |
| **21** | limit 19 to spanish |
| **22** | 20 or 21 |
| **23** | limit 22 to yr="2000 - 2022" |

#### Search strategy OVID MEDLINE^®^

Search strategy OVID MEDLINE ®
| # | Searches |
| --- | --- |
| **1** | exp multimorbidity/ |
| **2** | (multimorbid* or "multi morbid*" or comorbid* or "co morbid*").ti,ab. |
| **3** | (condition* adj3 count*).ti,ab. |
| **4** | ("multi condition*" or "multi disease*" or "multi disorder*" or multicondition* or multidisorder* or multidisease*).ti,ab |
| **5** | or/1-4 |
| **6** | exp "Transients and Migrants"/ |
| **7** | Refugees/ |
| **8** | exp "Emigrants and Immigrants"/ |
| **9** | (migrant* or refugee* or immigrant* or transient*).ti,ab. |
| **10** | (displace* adj3 person*).ti,ab. |
| **11** | (asylum* adj3 seeker*).ti,ab. |
| **12** | or/6-11 |
| **13** | exp Mortality/ |
| **14** | exp Death/ |
| **15** | prevalence/ |
| **16** | ((death* or fatalit* or mortalit*) adj3 (rate* or declin* or determinant* or "differential exces*")).ti,ab. |
| **17** | prevalence*.ti,ab. |
| **18** | or/13-17 |
| **19** | 5 and 12 and 18 |
| **20** | limit 19 to english |
| **21** | limit 19 to spanish |
| **22** | 20 or 21 |
| **23** | limit 22 to yr="2000 - 2022" |

#### Search strategy SCOPUS

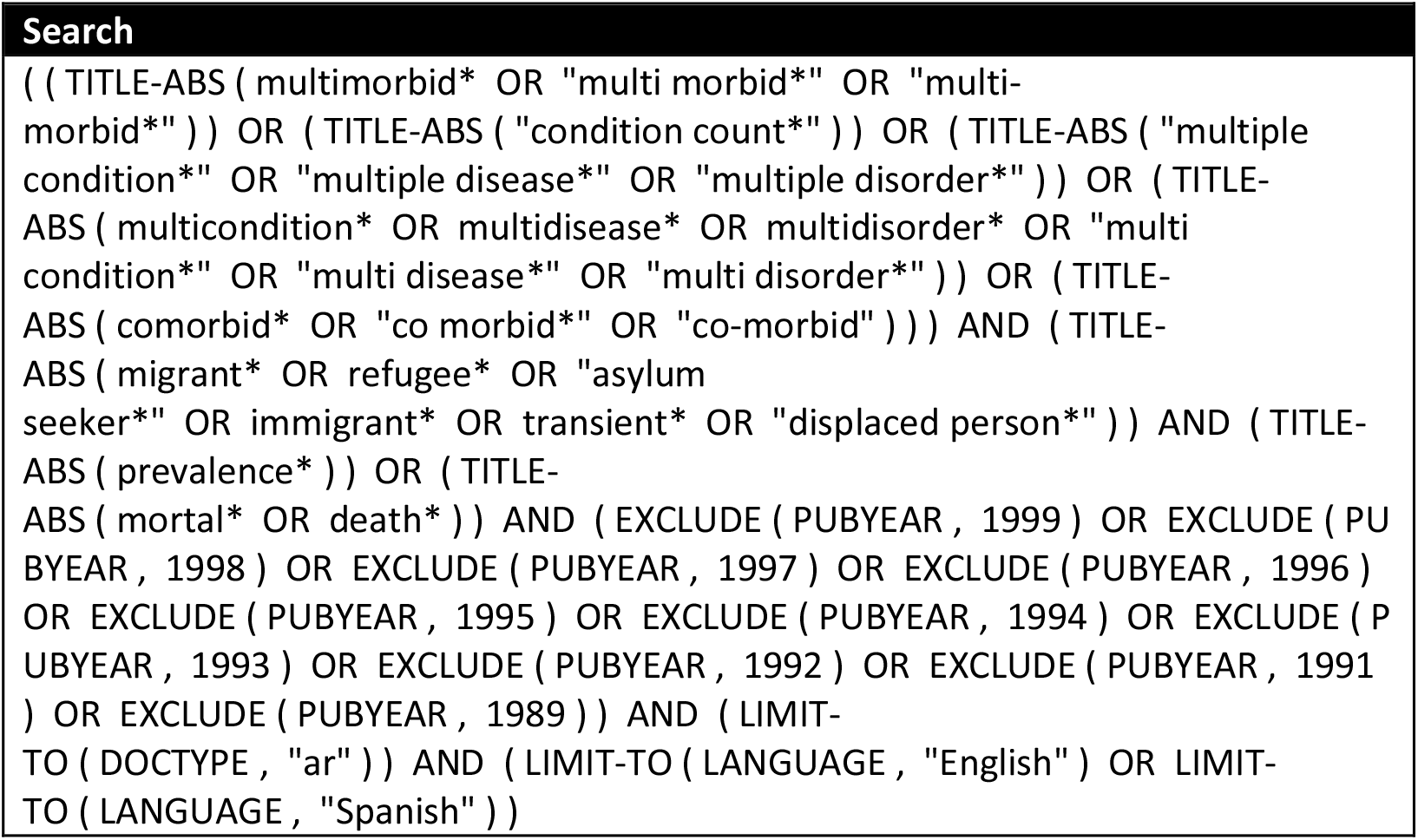

